# Prediction of the COVID-19 outbreak based on a realistic stochastic model

**DOI:** 10.1101/2020.03.10.20033803

**Authors:** Yuan Zhang, Chong You, Zhenghao Cai, Jiarui Sun, Wenjie Hu, Xiao-Hua Zhou

## Abstract

The current outbreak of coronavirus disease 2019 (COVID-19) has become a global crisis due to its quick and wide spread over the world. A good understanding of the dynamic of the disease would greatly enhance the control and prevention of COVID-19. However, to the best of our knowledge, the unique features of the outbreak have limited the applications of all existing models. In this paper, a novel stochastic model is proposed which aims to account for the unique transmission dynamics of COVID-19 and capture the effects of intervention measures implemented in Mainland China. We find that, (1) instead of aberration, there is a remarkable amount of asymptomatic individuals, (2) an individual with symptoms is approximately twice more likely to pass the disease to others than that of an asymptomatic patient, (3) the transmission rate has reduced significantly since the implementation of control measures in Mainland China, (4) it is expected that the epidemic outbreak would be contained by early March in the the selected provinces and cities.

## 1 Introduction

The current outbreak of coronavirus disease 2019 (COVID-19) has become a global crisis due to its quick and wide spread over the world. According to Official Report by the National Heath Commission of the People’s Republic of China (2020b), as of March 4, 2020, the outbreak of COVID-19 has caused 80 409 confirmed cases and 3012 fatalities in Mainland China. For the purpose of control and prevention, various containment measures have been implemented by the Chinese authorities, including traffic restrictions, medical tracking, entry or exit screening, isolation, quarantine and awareness campaigns since January 19, 2020. Especially on January 23, 2020, a strict travel restriction was introduced in Hubei province, cities in Hubei have been locked down since then (Government of the People’s Republic of China (2020)).

A good understanding of the epidemic dynamic would greatly enhance the control and prevention of COVID-19 as well as other infectious diseases, while dynamic model is probably one of the oldest and best-known mathematical tools to study the law of epidemic development. To the best of our knowledge, the SIR model proposed by Kermack and McKendrick (1927) almost a century ago is the first (and one of the most important) attempt to describe the development of an epidemic with a deterministic ordinary differential equation (ODE) system. Over the years, numerous modifications and generalizations have been made on the original SIR model to accommodate different epidemic characteristics, among which, the SIS model and SEIR model are most widely accepted ones. It is not surprising various deterministic dynamic models have been again designed and employed to the current outbreak of COVID-19. We hereby present a brief review on some of the representative works as follows.

Recently Tang et al. (2020) proposed a deterministic compartmental model by taking the clinical progression, epidemiological status, and the intervention measures into account. However, it implicitly assumed that the exposed population were not infectious, which is not the case in COVID-19. In addition, it was assumed that quarantine was implemented as soon as the infection occurred, which fails to reflect the inevitable latency brought by medical tracking. In the study of Wu et al. (2020), it proposed an extended SEIR model by considering transmissions among cities. However, it did not take the control measures such as quarantine and medical tracking into account. Furthermore, it was also assumed that exposed population are not infectious. For more similar or simpler deterministic ODE models to COVID-19, we refer to Liu et al. (2020b) for an overview.

Besides the ODE models, an even older approach is to discretize the time and consider a difference equation (DE) model, which can be seen as a self-propelled sequence of numbers (or vectors). In such models, the increment or decrement of different compartment at each step is a function of the configuration in the previous step(s). Earliest examples of a DE include the well-known geometric and Fibonacci sequence, which can both be used to describe the growth of a biological population. Recently, a discrete time DE model was introduced in Yang et al. (2020) to predict the epidemic trend of COVID-19. The proposed model correctly took the infectious incubation into account. However, this model did not consider the time lag between symptoms onset and diagnosis or the medical tracking. Besides, the rationale behind the assumption of the equal transmission probability between symptomatic and asymptomatic patients is questionable.

In contrast to the deterministic models (ODE or DE) summarized above, the transmission of a real world disease is inevitably random in nature. As a result, numerous stochastic dynamics models (see Section 3 for references) have been developed since the pioneering randomization of SIR model in Kendall (1956). In fact, a deterministic ODE model can often be seen as the mean-field equation of the corresponding stochastic counterpart. Under certain conditions, the mean-field equation may represent the evolution of the expectation of the corresponding stochastic model. In some more generalized cases, the mean-field equation is a large scale approximation of the corresponding stochastic model, which can be seen as a process version of *Law of Large Numbers*. However, when the size of outbreak is not comparable to that of the total population, the approximation aforementioned may not be interpreted that the stochastic and deterministic models are close to each other by themselves, since they have both been rescaled to approximately a constant in these scenarios. See Section 3 for more details. Thus compared with deterministic models, the stochastic model may be a better choice to account for the non-negligible random nature of the COVID-19 outbreak. Furthermore, the stochastic dynamic model is also known for its expandability to incorporate individual variations Athreya and Ney (1972), or even spatial structures Durrett (1988), which may not be fully captured by its mean-field equations. To our knowledge, the stochastic dynamic modeling for COVID-19 is yet relatively rare comparing to its deterministic counterparts, though preliminary approaches such as statistic exponential growth models was considered in recent studies of Liu et al. (2020a); Zhao et al. (2020). Very recently in Chinazzi et al. (2020), an existing discrete time, stochastic model (Balcan et al. (2009)) was employed to estimate the “effect of travel restrictions on the spread” of COVID-19. However, unique features of SARS-CoV-2, such as the infectious incubation and asymptomatic carriers, as well as control measures such as medical tracking, are still yet to be captured in Chinazzi et al. (2020).

To remedy the aforementioned issues in the existing studies, and to depict a more realistic transmission mechanism, we propose a novel stochastic compartmental model which captures the unique transmission dynamics of COVID-19 and the effects of intervention measures implemented in Mainland China. Our proposed stochastic model aims to study the COVID-19 outbreak in the following aspects:

- Estimation of key epidemiology parameters: Parameters in the model have specific epidemiology meanings. For example, *ρ* stands for the proportion of symptomatic virus carriers, *θ*_*E*_ represents the proportion of infectivity of asymptomatic carriers and symptomatic carriers and q can reflect the strength of quarantine in each region. These parameters have very limited information in medical reports for direct access and can be estimated through our model.
- Prediction of epidemic development: With a set of estimated parameters and initial values of the stages, we are able to simulate the development of epidemic over time t. This is helpful for capturing the spread tendency of the epidemic.
- Estimation of un-observable carriers and epidemic containment date: Un-observable virus carriers in the crowd are nonnegligible for assess of epidemic control. Instead of epidemic control assessment only through existing infections, our model is able to take the trend of un-observable carriers into account. Based on the simulated trend of un-observable carriers, epidemic containment date can also be calculated.
- Assessment of control measures The controlled reproduction number, *R*_*c*_, which reflects the transmission ability of the epidemic, is essential for the assessment of control measures and speed of disease transmission. *R*_*c*_ can be estimated from the ratio between the in-and-out flows of the active virus carriers in a given time period in our model. Moreover, we can also conduct hypothetical “controlled” test and evaluate the effectiveness of medical tracking.

Our main findings are as follows:

1. There exist a non-negligible portion of asymptomatic virus carriers.
2. Patients with symptoms have approximately twice the potential of transmission comparing with that of asymptomatic carriers.
3. The epidemic containment times in the selected provinces and cities (see Section 5 for a precise definition of the epidemic containment times) are estimated to be around late February to early March.
4. The time dependent controlled reproduction numbers *R*_*c*_ are between 2 and 3 in the selected provinces and cites at the beginning of the epidemic, and have been significantly reduced since the implement of control measures.
5. Medical tracking endeavors contribute significantly in containing the epidemic in the selected provinces and cities.

The rest of this paper is structured as follows. In Section 2 we describe the data used in this study. Section 3 introduces the proposed stochastic dynamic model and compares its features with some representative models in the recent literature. Parameter estimations are described in Section 4. In Section 5 we present our findings. We discuss our results, advantages and limitations in Section 6.

## 2 Method

### 2.1 Data Sources

Data used in this study include the numbers of confirmed diagnosis, recoveries and fatalities in the following provinces and cities of China: Beijing, Shanghai, Chongqing, Guangdong, Zhejiang and Hunan. These publicly available data were retrieved from Beijing Municipal Health Commission (2020); Shanghai Municipal Health Commission (2020); Chongqing Municipal Health Commission (2020); Health Commission of Guangdong Province (2020); Health Commission of Zhejiang Province (2020) and Health Commission of Hunan Province (2020) based on a daily update (see Table 5 and Table 6 in Appendix A). The corresponding population of residents in each region is collected from China National Bureau of Statistics (2018) (see Table 7 in Appendix A).

**Table 1:**
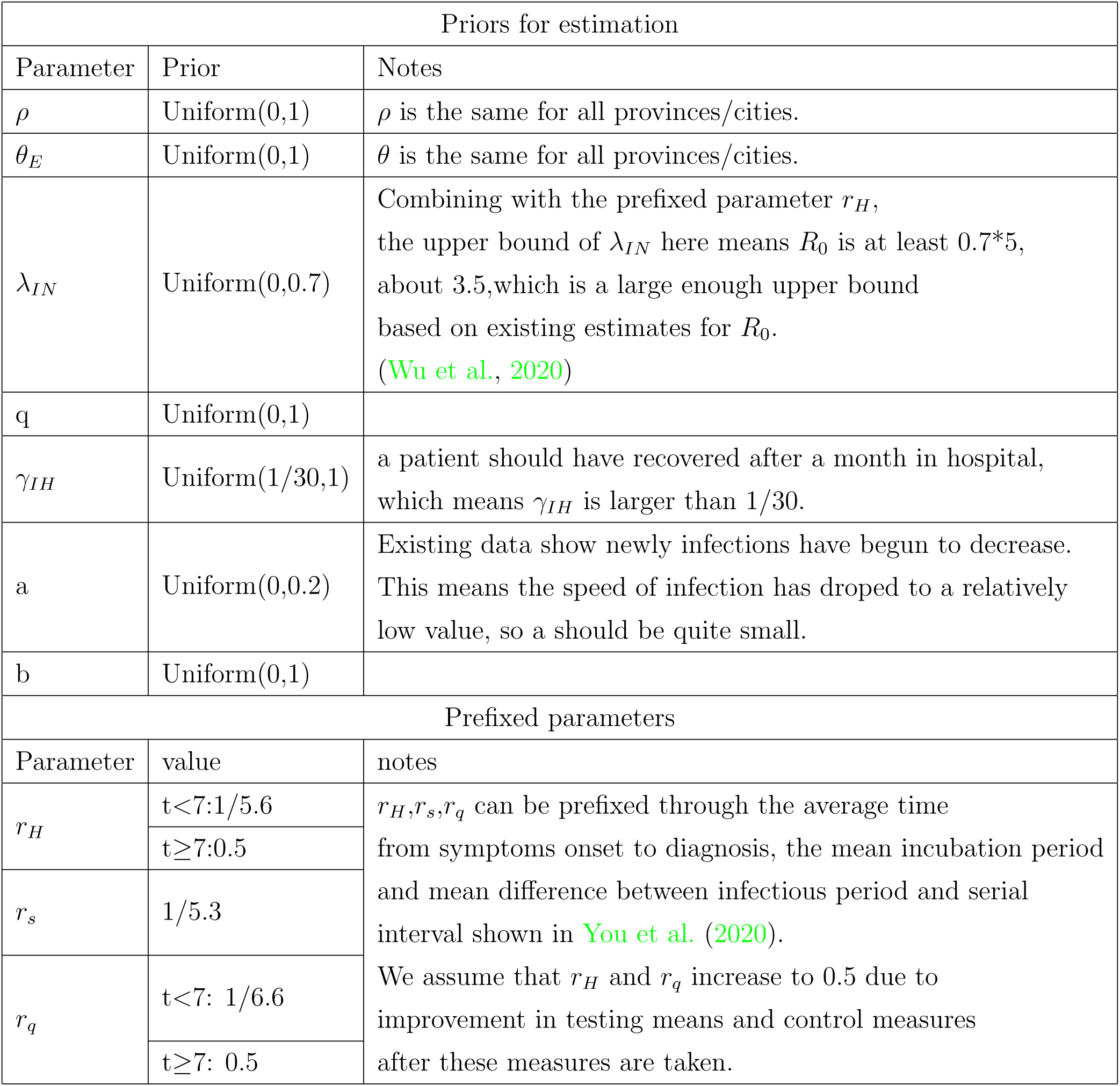
Priors for parameter estimation and values for prefixed parameters

**Table 2:** Initial Values

| Initial values |  |  |  |
| --- | --- | --- | --- |
| City | Parameter | Prior | Notes |
| Beijing | $E(0)$ | Uniform(0,179) | Upper bound for $IN(0)$ is obtained from the newly confirmed cases in the first 5 days.<br>Upper bound for $E(0)$ is obtained from a coarse estimate based on a conservative $R_0$ shown in Wu et al. (2020). |
| | $IN(0)$ | Uniform(0,66) | |
| Shanghai | $E(0)$ | Uniform(0,136) | |
| | $IN(0)$ | Uniform(0,50) | |
| Guangdong | $E(0)$ | Uniform(0,424) | |
| | $IN(0)$ | Uniform(0,156) | |
| Zhejiang | $E(0)$ | Uniform(0,443) | |
| | $IN(0)$ | Uniform(0,163) | |
| Chongqing | $E(0)$ | Uniform(0,275) | |
| | $IN(0)$ | Uniform(0,101) | |
| Hunan | $E(0)$ | Uniform(0,364) | |
| | $IN(0)$ | Uniform(0,134) | |

**Table 3:**
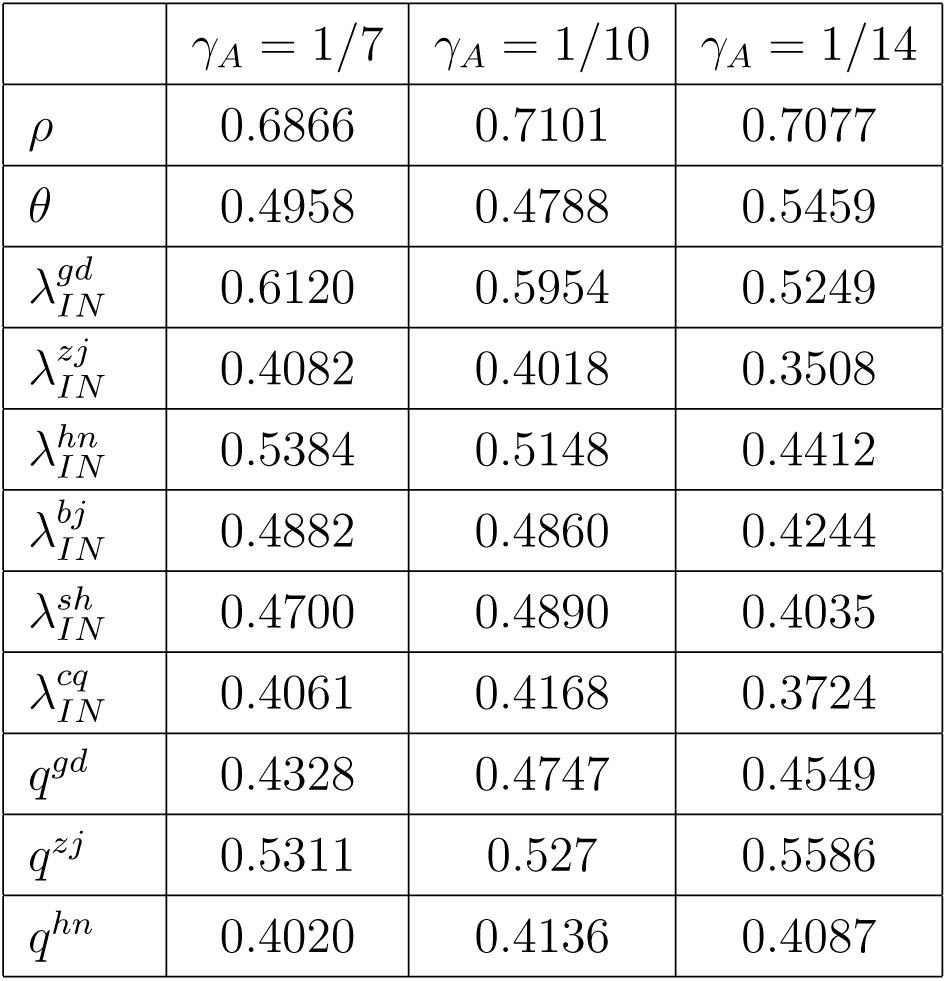
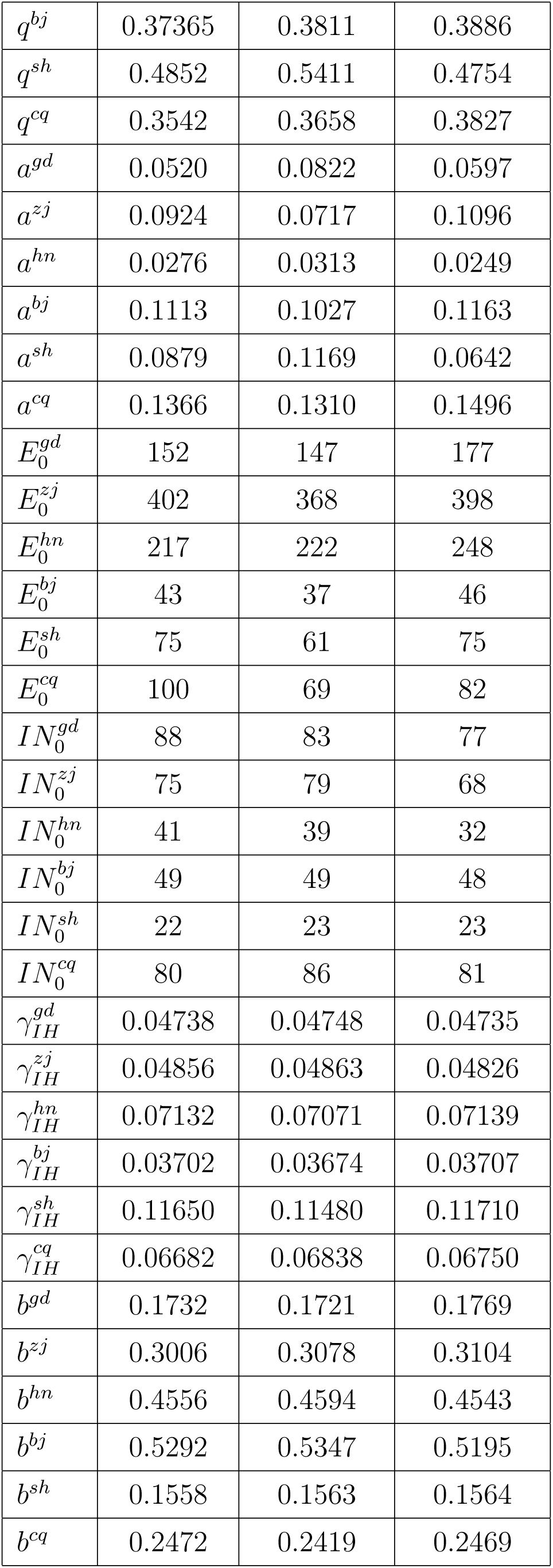
parameter estimation

**Table 4:** Comparison of Models

|  | Our Model | Tang et al. (2020) | Wu et al. (2020) | Yang et al. (2020) |
| --- | --- | --- | --- | --- |
| Model Type | Stochastic | ODE | ODE (with migration) | DE |
| Infectious Incubation | YES | NO | NO | YES |
| Asymptomatic Carrier | YES | YES | NO | NO |
| Medical Tracking | YES | YES | NO | NO |
| Latency MT* | YES | NO | NA | NA |
| Latency H** | YES | YES | NA | NO |

\* Latency MT= Latency in medical tracking
\*\* Latency H= Latency in hospitalization

Note that we exclude Hubei province which is the epicenter of the current outbreak in this study due to the following reasons: (1) the medical resources in Hubei province were overburdened at the beginning of the epidemic, not all individual with confirmed diagnosis could get immediate hospitalization; in fact according to the Official Press Briefing by the Information Office of Hubei Provincial People’s Government (2020), as of February 8, 2020, there were 1499 patients remained with confirmed diagnosis and serve symptom who had yet been hospitalized; (2) the diagnostic criteria were changed overtime in Hubei which resulted in a massive surge of confirmed cases in mid February (National Heath Commission of the People’s Republic of China (2020a)); (3) the fatality rate in Hubei province was much higher than other regions in China. These features distinct the dynamic model in Hubei from the model in other regions of China, which will be considered in our future studies, see Section 6 for details.

### 2.2 Model Description

The evolution of an epidemic is usually described by either a deterministic dynamic model Kermack and McKendrick (1927) or a stochastic one Kendall (1956); Bailey (1975), among which, the stochastic model is generally considered more realistic than the deterministic counterpart. Stochastic models often use a continuous time, compartment Markov Process to describe the evolution of the epidemic. See for instances, Kendall (1956); Kurtz (1970); Barbour (1972); Bailey (1975) for pioneering works, and Clémençon et al. (2008); Zhang and Wang (2013); Ji and Jiang (2014); Berrhazi et al. (2017) for recent developments. See also Wilkinson et al. (2017) and the references therein for a brief review.

Especially, when the size of outbreak is not comparable to that of the total population, the randomness is more significant, hence a stochastic model is needed to quantify the uncertainty in estimates and predictions in such case. In our study, none of selected provinces and cities has more than 2000 accumulated confirmed by now (see table 5 in Appendix A). These number, though alerting, are not comparable to the total population in provinces or cities, which are of an order 10 million-100 million (see table 7 in Appendix A). Hence, a novel stochastic dynamic model is designed to capture the unique features of the COVID-19 outbreak, where the **unique features** here refer to

- Infectious incubation period: according to an Official Press Briefing by the State Council Information Office of the People’s Republic of China (2020), unlike the SARS-CoV, SARS-Cov-2 can induce an infectious incubation period.
- Large portion of asymptomatic virus carriers: according to an Official Report by the Japanese Ministry of Health, Labour and Welfare (2020), it has been found that the proportion of asymptomatic infected population is considerable.
- Unprecedented contact control and medical tracking measures: since January 19, 2020, various containment measures have been implemented by the Chinese authorities. Especially on January 23, 2020, strict travel restriction was introduced in Hubei province at a unprecedented scale, cities in Hubei have been locked down since then (Government of the People’s Republic of China (2020)). At the same time, great efforts have also been taken in the medical tracking and quarantine of close contacts. For instance, remarkable efforts have been made by the The People’s Government of Zhejiang Province (2020), which till March 2, 2020 have successfully tracked more than forty thousand close contacts (People’s Government of Zhejiang Province (2020)).

To our knowledge, these features have not yet been fully captured by the existing stochastic dynamic models for the epidemic.

Under mild assumptions that (1) motions of all individuals in the system are independent, (2) the total population in the system is a fixed number of *N*, we devise the stochastic model with states listed as follow,

- *S*: Susceptible.
- *E*: Exposed. It is divided into four sub-states:
  – *E*_1_: will become symptomatic in the future and is not traceable in medical tracking.
  – *E*_2_: will become symptomatic in the future and is traceable in medical tracking.
  – *A*_1_: won’t become symptomatic in the future and is not traceable in medical tracking.
  – *A*_2_: won’t become symptomatic in the future and is traceable in medical tracking.

- *Q*: Quarantined. It is divided into two sub-states:
  – *E*_*q*_: will become symptomatic in the future.
  – *A*_*q*_: won’t become symptomatic in the future.
- IN: Infected, symptomatic, but not yet admitted to hospital. They are divided into two sub-states:
  – *IN*_1_: not traceable in medical tracking.
  – *IN*_2_: traceable in medical tracking.
- IH: Infected, symptomatic and currently under hospitalization. They are divided into two sub-states:
  – *IH*_*L*_: with light symptom.
  – *IH*_*S*_: with severe symptom.
- R: Recovered. They are divided into three sub-states:
  – *R*_*A*_: recover from state *A*_1_ and *A*_2_.
  – *R*_*N*_ : recover from state *IN*_1_ and *IN*_2_.
  – *R*_*H*_ : recover from state *IH*_*L*_.

- D: Dead.

Note that each individual can be classified into one of the above states at a specific time. For the purpose of conciseness, we denote *S*(*t*), *E*_1_(*t*), *E*_2_(*t*) and so on as the population sizes in the corresponding states at time *t*. The evolution of *ξ*(*t*) = {*S*(*t*), *E*_1_(*t*), *E*_2_(*t*), *E*_*q*_(*t*), *A*_1_(*t*), *A*_2_(*t*), *A*_*q*_(*t*), *IN*_1_(*t*), *IN*_2_(*t*), *IH*_*L*_(*t*), *IH*_*S*_(*t*), *R*_*A*_(*t*), *R*_*N*_ (*t*), *R*_*H*_ (*t*), *D*(*t*)} over time *t* forms a continuous time Markov Process with state space {0, _1_, 2, …, *N}*^15^, with transitions described as follows:

- Infection: Every primary case (including those of states *E*_1_, *E*_2_, *A*_1_, *A*_2_, *IN*_1_ and *IN*_2_) passes a pathogen to its secondary case at Poisson rate *λ*_*E*_ = *λ*_*IN*_ *θ*_*E*_, *λ*_*A*_ = *λ*_*IN*_ *θ*_*A*_ and *λ*_*IN*_ respectively. To be specific, a primary case chooses an individual randomly from the total population, and the individual is infected if it is of state *S*. At each transmission event,
  – with probability *ρ*, the secondary case becomes *E*_1_ or *E*_2_, meanwhile, this contact is traceable with probability *q*;
  – with probability 1 − *ρ* the secondary case becomes *A*_1_ or *A*_2_, meanwhile, this contact is traceable with probability *q*.

- Quarantine: Every individual of state *E*_2_, *A*_2_ and *IN*_2_ will be quarantined and consequently transfer into *E*_*q*_, *A*_*q*_ and *IH* respectively at a Poisson rate of *r*_*q*_. Note we assume such individuals will keep quarantined or will be hospitalized until recovery or death.
- Symptom onset: Every individual of state *E*_1_, *E*_2_ or *E*_*q*_ will develop symptoms and transfer into *IN*_1_, *IN*_2_ and *IH* respectively at a rate of *r*_*s*_.
- Hospitalization: Every individual of state *IN* will admitted to hospital at a Poisson rate of *r*_*H*_. With probability *p*_1_, it will transfer into *IH*_*L*_ and with probability 1−*p*1it will transfer into *IH*_*S*_.
- Symptom relief: Every individual of state *IH*_*S*_ will transfer into *IH*_*L*_ at a Poisson rate of *r*_*b*_.
- Recovery: Every individual of state *A*_1_, *A*_2_, *IN* and *IH*_*L*_ will recover at Poisson rates of *γ*_*A*_, *γ*_*IN*_ and *γ*_*IH*_ respectively.
- Death: Every individual of state *IN* and *IH*_*S*_ will die at Poisson rates of *δ*_*IN*_ and *δ*_*IH*_ respectively.

The process can be illustrated by Figure 1. The precise definition of the continuous time Markov process can be found in Appendix B.

**Figure 1:**
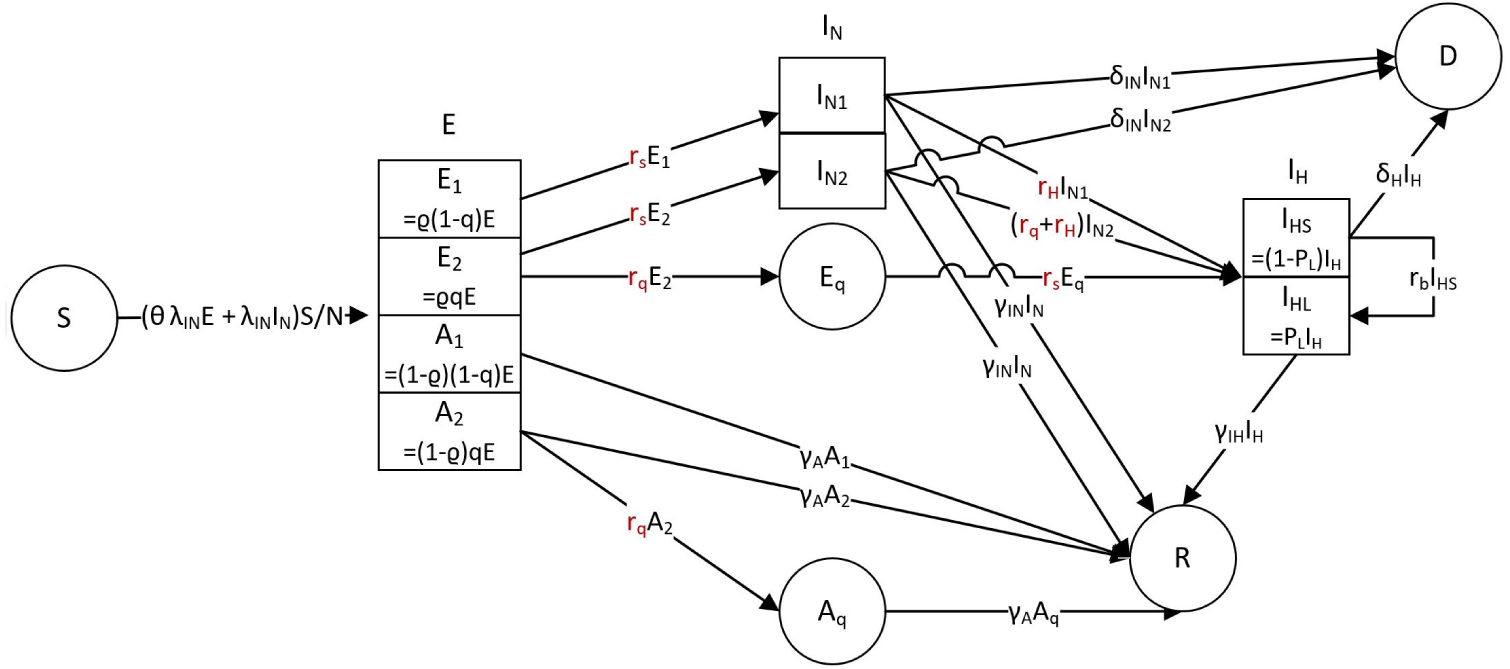
Process Illustration

From a more theoretical, functional analysis point of view, the stochastic dynamic model defined above can be equivalently described with its Markov Semigroup/infinitesimal generator, see Liggett (1985); Ethier and Kurtz (1986) for details. From those operators, it is natural for us to consider the following **Mean-field Differential Equation System**, which itself serves as a deterministic counterpart of the stochastic model, that is,

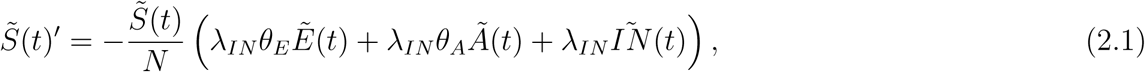

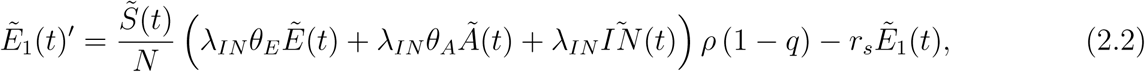

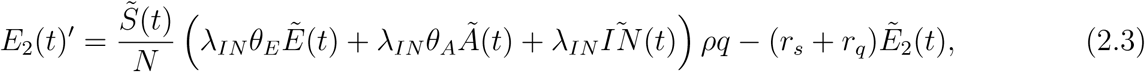

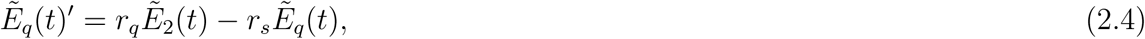

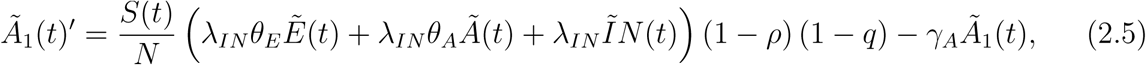

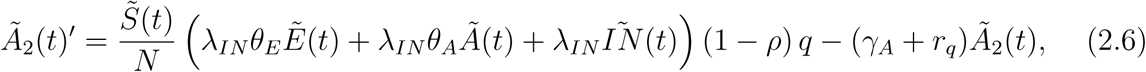

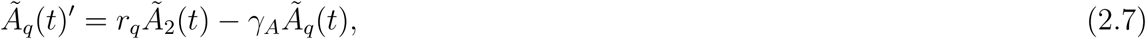

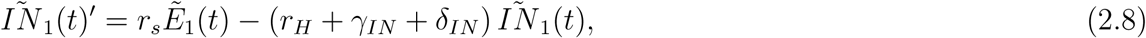

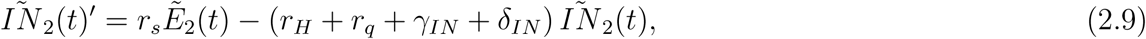

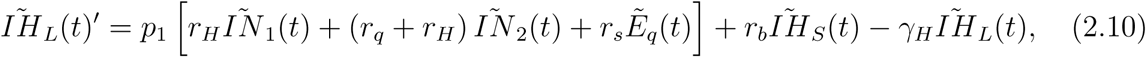

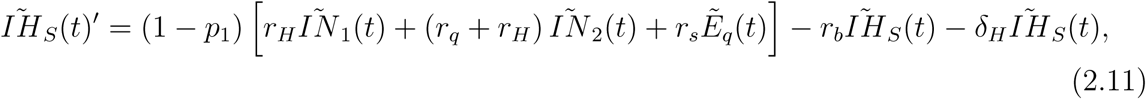

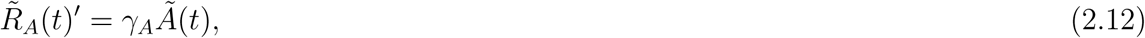

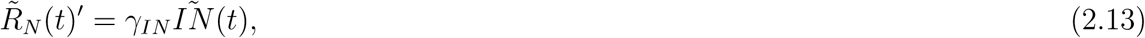

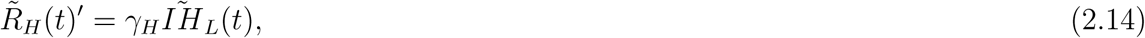

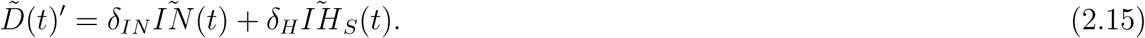

Intuitively, the mean-field ODE system above serves as a degenerated case of our stochastic model, where all randomness has been averaged out. When the differential equations are linear, which is not true in our case, the ODE also describes the evolution of the expectation of the stochastic system (Dynkin’s Formula, see Da Prato and Zabczyk (2014) Section 4.4 for example). Besides, according to Kurtz (1970) and Kurtz (1971), we may see that this deterministic model can also serve, under a much weaker condition, as a scaling approximation of the stochastic one after recaled by the renormalizing factor *N*. To be more specific, the renormalized random path *ξ*_*N*_ (*t*)*/N* as a stochastic process is “almost” deterministic, and fluctuates closely around the deterministic trajectory 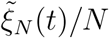 if the total population *N* is large and the renormalized initial values *ξ*_*N*_ (0)*/N* and 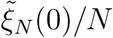 are close to each other, where 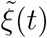 denote the collections of 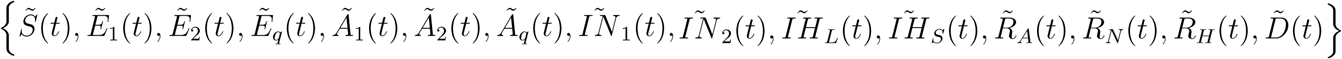 and 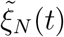 and *ξ*_*N*_ (*t*) be copies of the deterministic and stochastic model with total population *N*. Using a mathematical language, with probability one there is the pathwise convergence

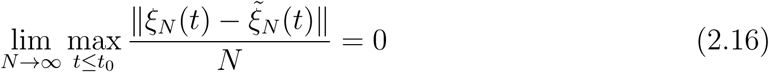

for all *t*_0_ ∈ [0, ∞). However, we would like to specifically bring to the reader’s attention that, NO MATTER the size of the total population *N*, as long as the size of epidemic outbreak is NOT comparable to *N*, the convergence above in (2.16) DOES NOT imply that the un-renormalized *ξ*(*t*) is non-random by itself or that the epidemic involved populations. For example, *E*_1_(*t*) and 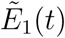 are close to each other in any sense without the rescaling factor. Actually the fact that the size of epidemic outbreak is not comparable to *N* itself already implies that

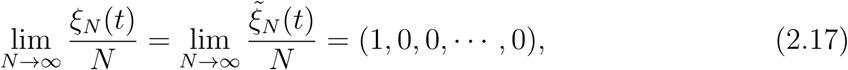

which perfectly satisfies (2.16) while at the same time provides no information on whether the actual values of *E*_1_(*t*) and 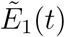 are close to each other. To summarize, when the size of epidemic outbreak is NOT comparable to *N*, the stochastic model possesses intrinsic randomness, and may not be well approximated by a deterministic ODE in any non-degenerate sense.

### 2.3 Choices of initial values and estimation of model parameters

The parameters and initial values of the proposed stochastic model can be estimated based on a simplified model in Figure 2. Here the stage *D* is removed as fatality rate is extremely low in all the selected regions. The populations of *IH*(*t*), *R*_*H*_ (*t*) can be observed directly from the collected data, that is, the number of existing confirmed cases and reported recoveries at time *t* respectively, while, the remaining states are latent, namely not observable. Among the latent states, the initial value *S*(0) can be approximated by the population of permanent residents in the city or province, *E*_*q*_(0) is set to be zero as there was no quarantine implemented before January 23, 2020 and *R*_*N*_ (0) can be set to any number as it would not affect the estimation and prediction of the model. However, the choices for initial values, *IN* (0) and *E*(0), is also non-observable and could be a challenge to determine apriori Peng et al. (2020). In this study, *IN* (0) and *E*(0) are treated as unknown parameters and to be estimated together with other model parameters as described below.

**Figure 2:**
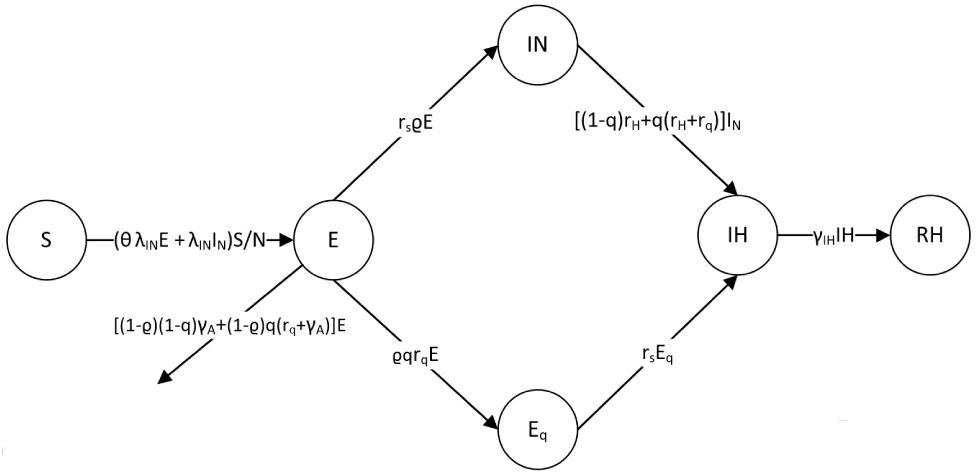
A Simplified Version of process

There is a total of 9 model parameters in the proposed model for each selected region. They are *λ*_*IN*_, *θ*_*E*_, *ρ, q, γ*_*IH*_, *γ*_*A*_, *r*_*s*_, *r*_*q*_ and *r*_*H*_, among which, *r*_*s*_, *r*_*q*_ and *r*_*H*_ are related to the clinical characteristics of the disease and can be prefixed through existing studies. To be more specific, *r*_*H*_ is the inverse of the average time from symptoms onset to diagnosis, *r*_*s*_ is the inverse of the mean incubation period, while *r*_*q*_ is the inverse of mean difference between infectious period and serial interval. Based on preliminary trials, we find there is very limited information of *γ*_*A*_ which can be obtained from the data, and the estimate is highly influenced by the choices of prior. A possible explanation towards it is that *γ*_*A*_ is less related to the observations. Hence, instead of estimating *γ*_*A*_ with large uncertainty, we prefix *γ*_*A*_ = 1*/*10. Sensitivity analysis is conducted on the different choices of *γ*_*A*_.

The rest of parameters would be estimated from the model. Among which, the parameters, *ρ* and *θ*_*E*_, are directly related to the nature of the disease, and hence are considered as constants in China, while, *λ*_*IN*_, *q* and *γ*_*IH*_ may vary in different regions depending the local medical resources, population densities and containment measures. Furthermore, it is more realistic to consider *λ*_*IN*_ and *γ*_*IH*_ as time varying parameters to reflect the effect of intervention measures and improvement of the medical treatment. In this study, a simple setting for the time varying function is used, that is, 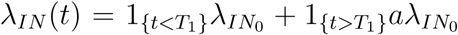 and 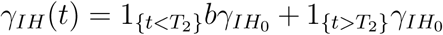. The time *T*_1_ is set to be January 29 as there is an obvious change of rates occurred on January 29 illustrated in Figure 3 (You et al., 2020). We use observed 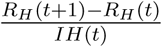 to approximate *γ*_*IH*_ on day *t*. The time *T*_2_ is selected to be the time when *γ*_*IH*_ has a significant change for each province/city.

**Figure 3:**
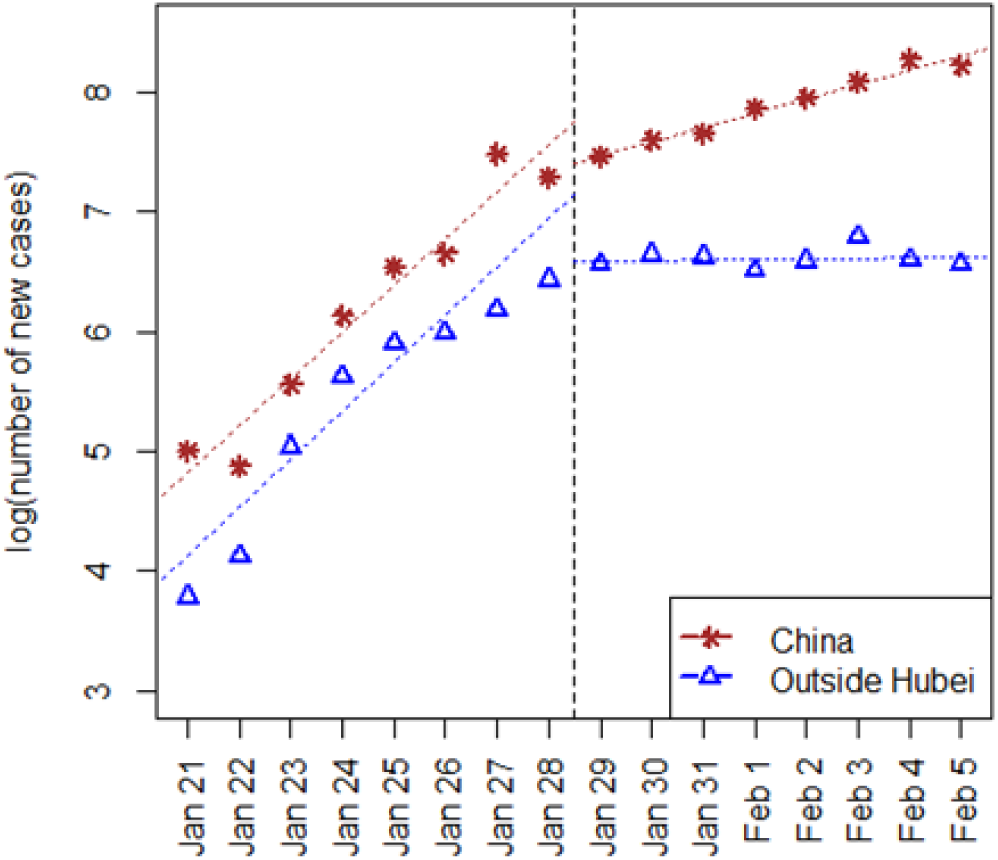
Visualization of daily numbers of newly confirmed case along with date, an obvious change of rate occurred on January 29, 2020.

The likelihood function is obtained by assuming that daily confirmed cases and recoveries are independent Poisson random variables (Wu et al., 2020), that is,

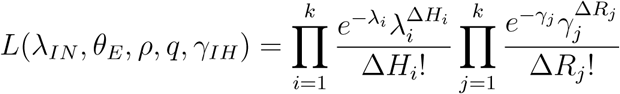

where Δ*H*_*t*_ and Δ*R*_*t*_ are the newly confirmed and recovered cases on day *t, λ*_*t*_ and *γ*_*t*_ are the functions of model parameters *E*(0), *IN* (0),*λ*_*IN*_, *θ*_*E*_, *ρ, q* and *γ*_*IH*_ based on the Mean-field Differential Equation System. Parameters are estimated by the posterior means through the Metropolis-Hastings algorithm implemented in the R package POMP (King et al., 2020, 2016) where non-informative uniform distributions are chosen as the prior distributions, see Table 1 and 2 for more details.

## 3 Results

### 3.1 Interpretation of parameters

A summary of the estimated model parameters is given in Table 3, from which we find that

- The estimate of *ρ* is not sensitive to the choice of *γ*_*A*_, about 30% infected individuals are asymptomatic.
- The estimate of *θ*_*E*_ is reasonably close to each other with different choices of *γ*_*A*_, patients with symptoms are about twice as likely to pass a pathogen to others as asymptomatic patients.
- The estimate of *q* is not sensitive to the choice of *γ*_*A*_ except in Shanghai. Zhejiang has the highest *q* in the selected regions, which is consistent with the remarkable efforts made by the The People’s Government of Zhejiang Province (2020), which till March 2, 2020 has successfully tracked more than forty thousand close contacts (People’s Government of Zhejiang Province (2020)).

The sensitive analysis of *γ*_*A*_ shows that, (1) the estimates of *γ*_*IH*_ and *b* are consistent regardless the choice of *γ*_*A*_ as the states *RH* and *IH* are both observable; (2) the estimated initial populations for states *E* and *IN* in each regions vary over different choices of *γ*_*A*_, but are still in the same order of magnitude; (3) the estimate of *λ*_*IN*_ and *a* vary with the choices of *γ*_*A*_ notably in Guangdong, Zhejiang and Shanghai, cautions need to be taken when making inferences on *λ*_*IN*_ and *a*.

### 3.2 Prediction of the epidemic trend

Based on the estimated parameters, future trajectories of the epidemic are simulated using the proposed stochastic dynamic model. For each region, 1000 simulations are conducted to produce the 95% confidence interval for the epidemic evolution of some key populations. In this section, the predictions of populations of states, the containment time of the outbreak, the controlled reproduction number *R*_*c*_, and a test on the effectiveness of the current medical tracking policy are reported.

Figure 4 plots the 95% confidence interval of:

- accumulated confirmed cases, namely, the sum of *IH, RH* and *D*.
- the population of state *IH*, representing the people in hospitals.
- population of active virus carriers, consisting of the states *E* and *IN*.

**Figure 4:**
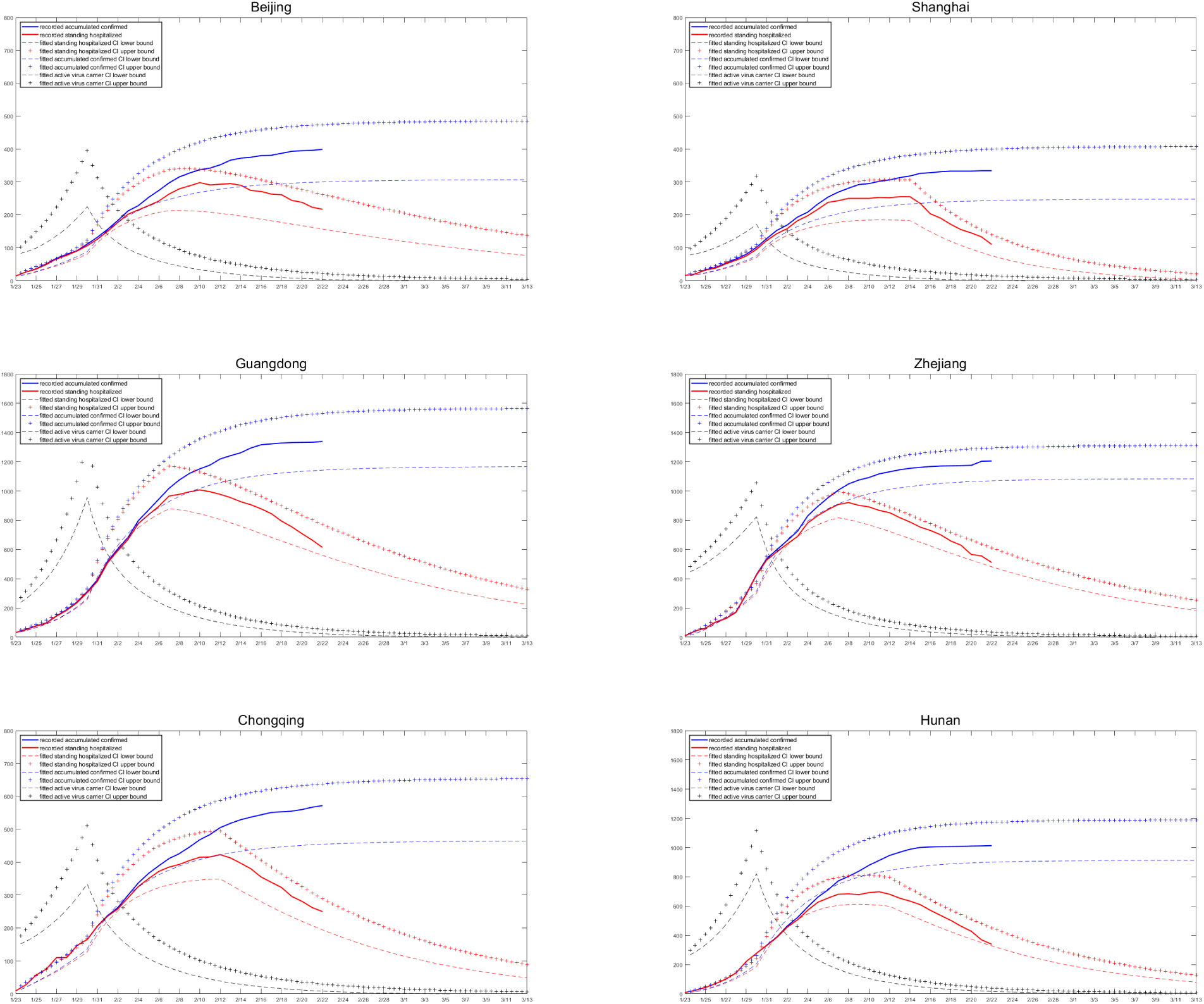
Predicted confidence interval for key populations

Note that the first two populations can be directly observed, but population of *E* and *IN* is not observable. We can see that the observed accumulated confirmed cases and the population in *IH* lie in the calculated confidence interval well. Sensitivity analysis shows that the predicted confidence interval is stable against the uncertainty of *γ*_*A*_, which gives us more confidence in that the proposed model can capture the spread tendency of the epidemic.

The containment time of the outbreak is defined as the moment when the number of active virus carriers is first less than a threshold *T*_*c*_. Noting that the spread of the disease is due to the existing active virus carriers in the proposed model, thus when the number of the active virus carriers decreases to a certain level, the new infections are limited, which means the epidemic is under control. Figure 5 shows the 95% confidence interval of the containment time of the outbreak for each region when *T*_*c*_ = 10. Among the six regions in this study, Shanghai is predicted to have the earliest containment time of February 21, while the containment time in Guangdong is predicted to be the latest, around March 7. From the daily report of NHC of China, new infections in Shanghai from February 17 to February 21 are 0,0,0,1,0; and new infections in Guangdong in the same period are 6,3,1,1,6, which potentially supports our prediction. Notice that in sensitivity analysis on *γ*_*A*_, we find that when the recover rate of asymptomatic patients *γ*_*A*_ gets smaller, the expected containment time is longer. This is because smaller *γ*_*A*_ means that infected asymptomatic patients need more time to recover, which making the number of active virus carriers stays large for a longer time.

**Figure 5:**
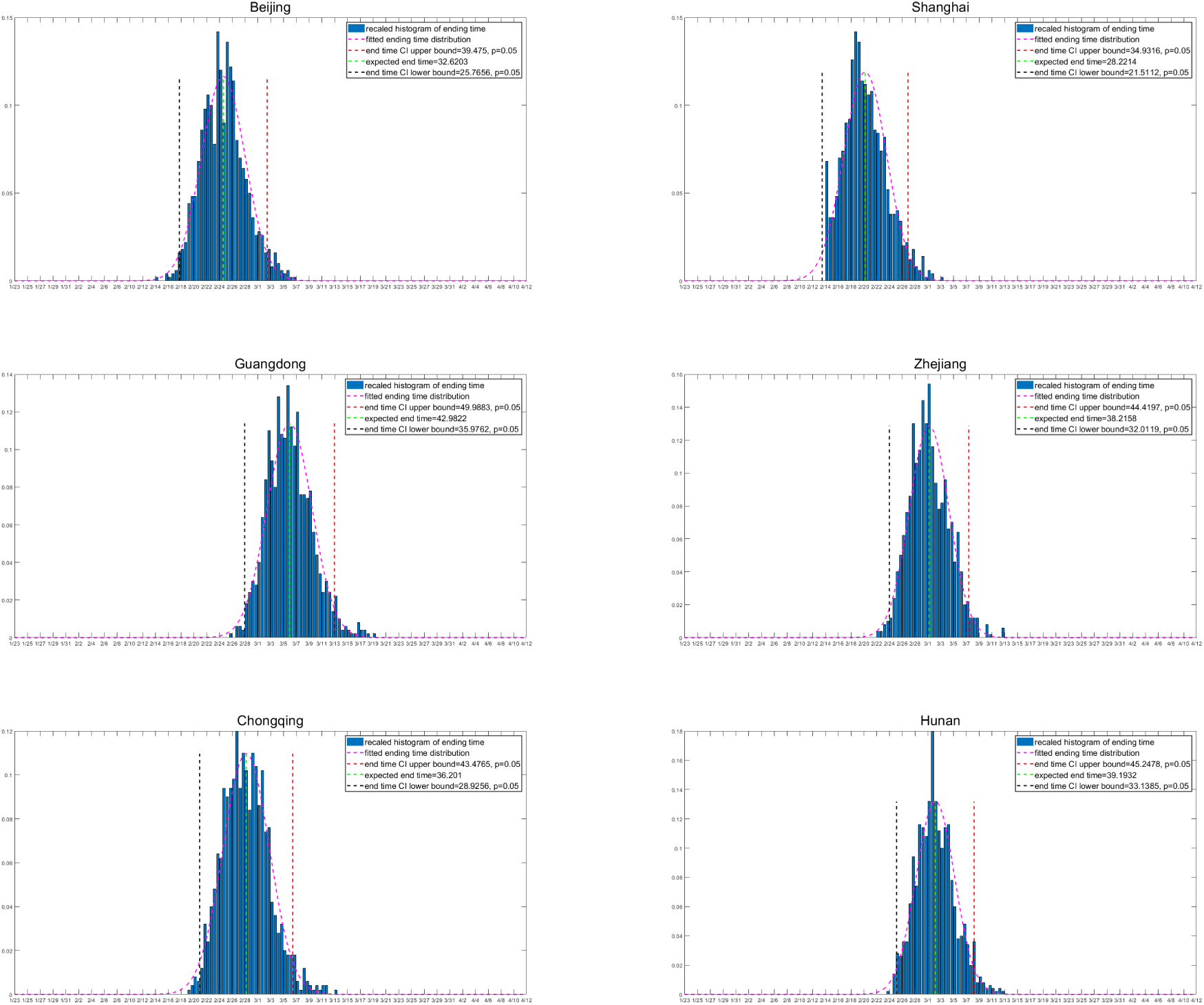
Prediction of the containment time of the outbreak. The containment time of 1000 simulations is plotted as a histogram and is fitted with normal distribution for each region. The *y* axis represents the density of containment time.

The controlled reproduction number, *R*_*c*_, reflects the transmission ability of the epidemic, which is one of the most important quantities in epidemiology. In this study, the evolution of *R*_*c*_ is approximated by the ratio between the in-and-out flows of the active virus carriers in a given time period, that is, for a time interval of length Δ*t*, say [*t, t* + Δ*t*], we keep tracking the transitions that leads to the increase/decrease of the active virus carrier population *E* and *IN*, with their accumulative numbers recorded. Precisely speaking, we define

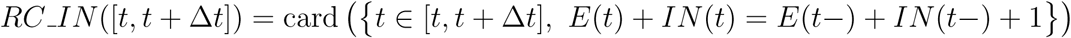

and

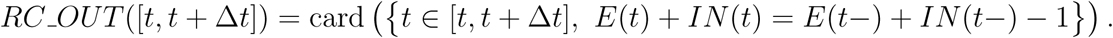

Recalling that our stochastic epidemic model evolves as a non-explosive continuous time Markov process, *RC_IN* (*·*) and *RC_OUT* (*·*) are both non-negative finite integers. And when *RC_OUT* ([*t, t* + Δ*t*]) ≠ 0, we approximate the controlled reproduction number over [*t, t* + Δ*t*] by its average value

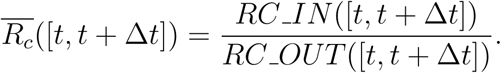

Simulation results for the approximated *R*_*c*_ in each region is shown in Figure 6. In most provinces and cities we find *R*_*c*_ is between 2 and 3 before control measures, and drops rapidly to about 0.2 during January 29 and February 1 in all selected regions.

**Figure 6:**
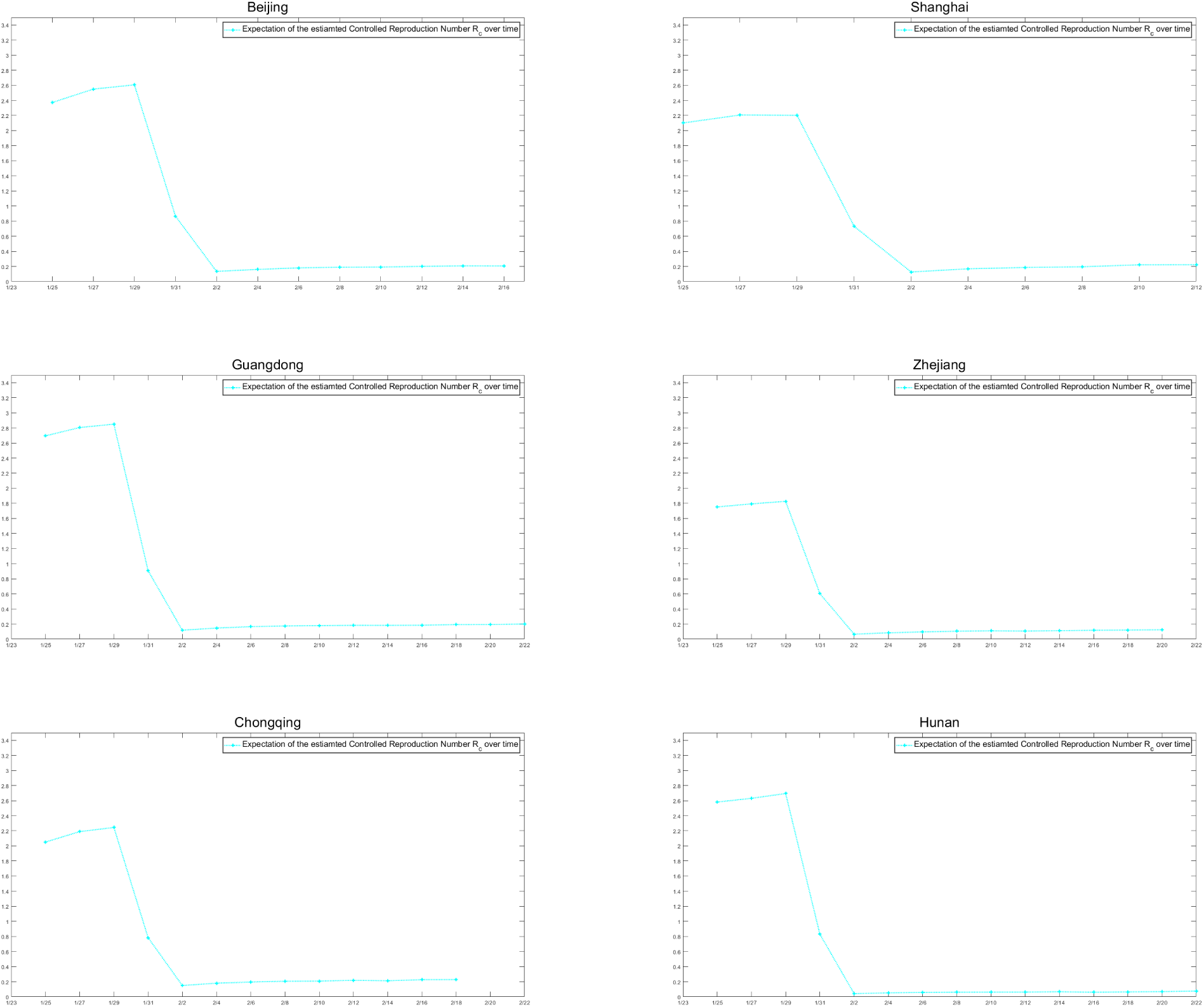
Predicted time-varying *R*_*c*_ curve.

Finally, we evaluate the effectiveness of the current medical tracking policy with a hypothetical controlled test as follows: while keeping the other parameters we estimated unchanged, we set the traceable ratio *q* = 0 to represent the scenario where no medical tracking were implemented, and let the system evolve. Under this setting, the epidemic would still be contained, see Figure 7 for details, due to the contact control measures that reduce the values of *λ*’s and the reduction of diagnosis waiting time. However, for selected the provinces and cities in this study, there are significant delays in the dates of containment when *q* = 0, which indicates the current medical tracking policy contribute significantly in containing the epidemic.

**Figure 7:**
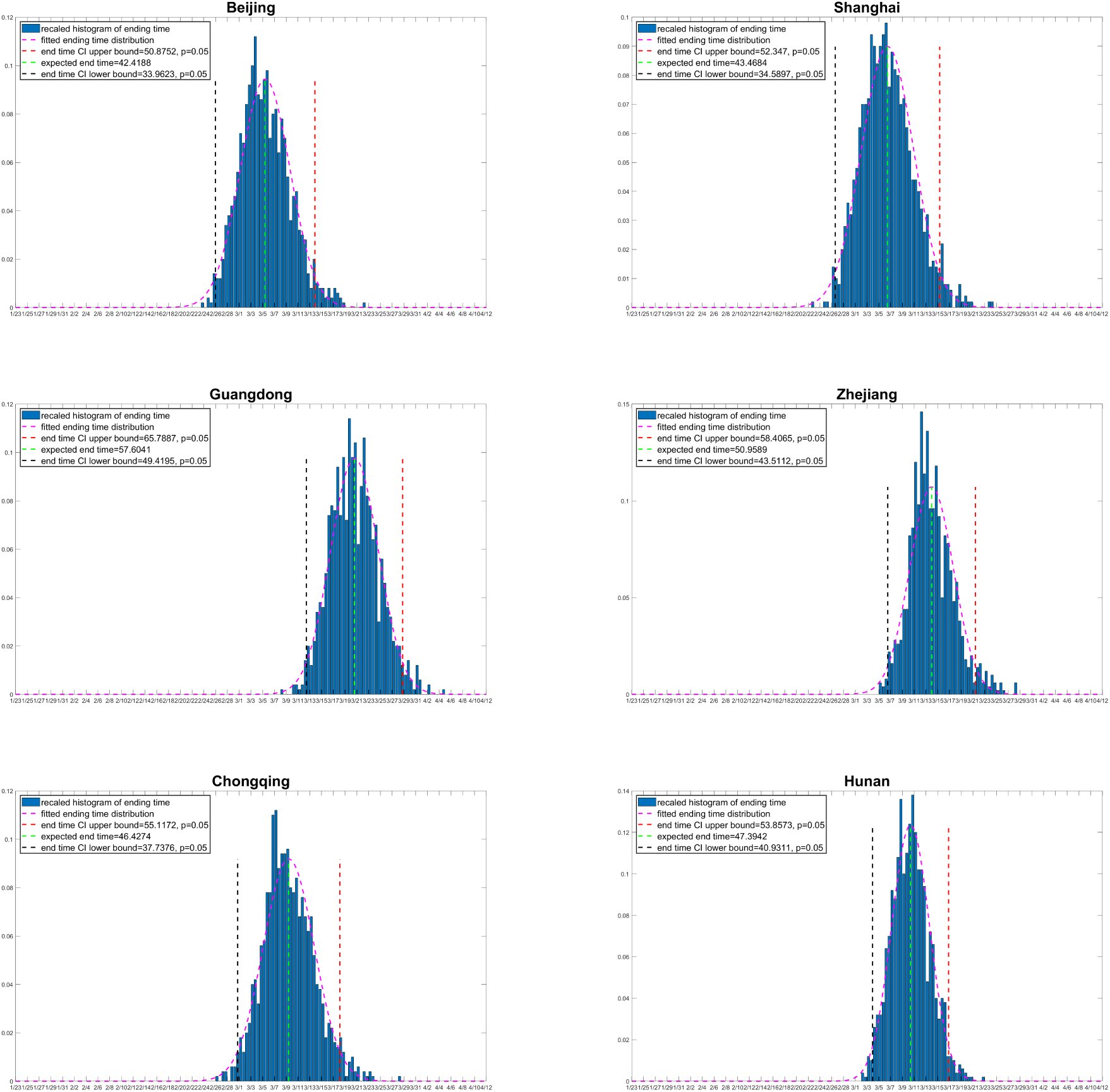
Prediction of the containment time of the outbreak with *q* = 0. The containment time of 1000 simulations is plotted as a histogram and is fitted with normal distribution for each region. The *y* axis represents the density of containment time.

## 4 Discussion

In this article, we propose a novel stochastic dynamic model in order to depict the transmission mechanism of COVID-19. In comparison with some existing models on COVID-19, our model features the employment of a stochastic dynamic as well as a comprehensive account for the infectious incubation period, the asymptomatic virus carriers, and the medical tracking measure with time latency, which make it probably closer to reality in these aspects. Moreover our proposed model also sets the basis for further studies with individual/network based models, which may not have an exact mean-field counterpart.

In the selected provinces and cities in our study, none of them has more than 2000 accumulated confirmed by now (See Table 5 in Appendix A). The numbers of confirmed cases, though alerting, are still not comparable to the total populations of these provinces, which are of the order 10 million-100 million (see Section 2 for exact population data). Moreover, for a system of size 1000 or so, it is not safe to argue the randomness is negligible due to the Law of Large Numbers, which makes it already sufficiently close to its hydrodynamic limit. This justifies the use of stochastic dynamic in our model.

Since the outbreak of COVID-19, the government has conducted large-scale medical tracking and quarantine measures. These non-negligible measures have a significant impact on the spread of COVID-19. The sub-states of *E, A* and *IN* in our model are able to reflect such tracking and quarantine measures. Moreover, we also take the latency between medical tracking and quarantine into account. What’s more, it has been found that the proportion of asymptomatic infected population is considerable. This justifies the introduction of state *A*. Finally, for an exposed and symptomatic individual, this model describes the complete process from infection to recovery(or death).

In comparison, the model of Tang et al. (2020)(hereafter abbreviated as T.model) failed to consider the infectivity of the incubative virus carriers(*E*). What’s more, T.model assumed quarantine measures will complete at the moment of contact. It is not completely physical since there should always be a time lag between the transition and the medical tracking. In Wu et al. (2020), they didn’t explicitly consider the impact of medical tracking and quarantine measures in dynamic. As we mentioned above, there are large-scale medical tracking and quarantine measures after the outbreak of COVID-19, and these measures have a significant impact on the spread of the virus. The questionable assumption that the exposed population is non-infectious has also been made in these two models.

The recent work of Yang et al. (2020) proposed a discrete time difference equation(DE) which correctly accounted the infectious incubation period. However, they assumed a symptomatic patient is hospitalized (“quarantine” in their term, see Page 4 of Yang et al. (2020)) immediately at symptom onset, and did not consider the time lag between symptom onset diagnosis/hospitalization. Moreover, the probability of transmission *b* are questionably assumed to be the same for both the symptomatic and asymptomatic population. See Page 3-4 of Yang et al. (2020); and no medical tracking mechanism has been considered. Finally, there seems to be multiple imperfections in the interpretation of dynamic and estimation of parameters.

The differences among models mentioned above are hereby summarized in Table 4:

From the estimated parameters, we find that about 30% of the infected are asymptomatic, patients with symptoms are about twice as likely to pass a pathogen to others as asymptomatic patients, and current containment measures are effective to reduce the contact and transmission rate. The containment time of the outbreak for different regions are predicted. Time-varying *R*_*c*_ is calculated and a rapid drop of *R*_*c*_ is observed due to containment measures that governments take. We have also conducted controlled test and find that in addition of the control measures reducing the contacts of people, the current medical tracking policy also contributes significantly in containing the epidemic.

However, we acknowledge there are limitations in the propose model,

1. Given the limited data available, certainly parameters, especially those “far away” from observation in the proposed model may have a potential risk of identification issues.
2. The current estimation method converges slowly;
3. The proposed model does not apply if significant changes apply to the current epidemic control/treatment measure in the future;
4. The proposed model need further modification if a non-negligible portion of the asymptomatic patients remain infectious after the end of quarantine.
5. Parameter estimates may lose precision if the stochastic model differs excessively from its simplification described in Section 4.

In our future study, we propose to complement/generalize our current stochastic model from the following aspects:

- **Improved medical tracking dynamic**: In this ongoing work, we are to introduce a more realistic dynamic for medical tracking. Our current model has already considered the latency between transmission and quarantine. While in this modification, medical tracking is triggered in a more physical manner by the affirmative diagnosis of the transmission source. In this new model the quarantine process of each exposed agent depends on his/her individual contact history. Thus an individual based, rather than compartment dynamic is needed. Then the parameters we have estimated can be used as a basis for the individual-based model.
- **Introduction of medical service capacity**: In this modification, the maximal capacity of the medical service system is to be considered, which might be overloaded when facing a massive outbreak. This idea was first inspired by an Amateur Demonstration made by Ele laboratory (2020) on a site named “bilibili”, and seems to be among the key features of the initial phase of the COVID-19 outbreak in Wuhan. We may also allow such capacity to be time/configuration dependent to model the contribution of building the “emergency hospitals” in Wuhan.
- **Population flows over cities**: We will model the migration of people over different regions, which could have played an essential role in the spread of the epidemic before the Chinese New Year of 2020. We will also consider the transmissions on board and even allow the population flow to react on the information they have about the epidemic situation.

## Data Availability

Data is attached in the appendix

## Acknowledgment

This research is supported by National Natural Science Foundation of China grant 8204100362 and Zhejiang University special scientific research fund for COVID-19 prevention and control.

## Appendix A: Data

**Table 5:** Culmulative confirmed cases. Data for the six regions is available at Beijing Municipal Health Commission (2020); Shanghai Municipal Health Commission (2020); Chongqing Municipal Health Commission (2020); Health Commission of Guangdong Province (2020); Health Commission of Zhejiang Province (2020); Health Commission of Hunan Province (2020).

| date | Beijing | Shanghai | Chongqing | Guangdong | Zhejiang | Hunan |
| --- | --- | --- | --- | --- | --- | --- |
| 2020.01.22 | 14 | 16 | 9 | 32 | 10 | 9 |
| 2020.01.23 | 26 | 20 | 27 | 53 | 43 | 24 |
| 2020.01.24 | 36 | 33 | 57 | 78 | 62 | 43 |
| 2020.01.25 | 51 | 40 | 75 | 98 | 104 | 69 |
| 2020.01.26 | 68 | 53 | 110 | 146 | 128 | 100 |
| 2020.01.27 | 80 | 66 | 110 | 188 | 173 | 143 |
| 2020.01.28 | 91 | 80 | 147 | 241 | 296 | 221 |
| 2020.01.29 | 111 | 101 | 165 | 311 | 428 | 277 |
| 2020.01.30 | 132 | 128 | 206 | 393 | 537 | 332 |
| 2020.01.31 | 156 | 153 | 238 | 520 | 599 | 389 |
| 2020.02.01 | 183 | 169 | 262 | 604 | 661 | 463 |
| 2020.02.02 | 212 | 193 | 300 | 683 | 724 | 521 |
| 2020.02.03 | 228 | 208 | 337 | 797 | 829 | 593 |
| 2020.02.04 | 253 | 233 | 366 | 870 | 895 | 661 |
| 2020.02.05 | 274 | 254 | 389 | 944 | 954 | 711 |
| 2020.02.06 | 297 | 269 | 411 | 1018 | 1006 | 772 |
| 2020.02.07 | 315 | 281 | 426 | 977 | 1048 | 803 |
| 2020.02.08 | 327 | 292 | 446 | 1120 | 1075 | 838 |
| 2020.02.09 | 337 | 295 | 468 | 1151 | 1092 | 879 |
| 2020.02.10 | 342 | 302 | 484 | 1177 | 1117 | 912 |
| 2020.02.11 | 352 | 306 | 505 | 1219 | 1131 | 946 |
| 2020.02.12 | 366 | 313 | 518 | 1241 | 1145 | 968 |
| 2020.02.13 | 372 | 318 | 529 | 1261 | 1155 | 988 |
| 2020.02.14 | 375 | 326 | 537 | 1294 | 1162 | 1001 |
| 2020.02.15 | 380 | 328 | 544 | 1316 | 1167 | 1004 |
| 2020.02.16 | 381 | 331 | 551 | 1322 | 1171 | 1006 |
| 2020.02.17 | 387 | 333 | 553 | 1328 | 1172 | 1007 |
| 2020.02.18 | 393 | 333 | 555 | 1331 | 1173 | 1008 |
| 2020.02.19 | 395 | 333 | 560 | 1332 | 1175 | 1010 |
| 2020.02.20 | 396 | 334 | 567 | 1333 | 1203 | 1011 |
| 2020.02.21 | 399 | 334 | 572 | 1339 | 1205 | 1013 |

**Table 6:** Cumulative recoveries. The data for the six regions are available at Beijing Municipal Health Commission (2020); Shanghai Municipal Health Commission (2020); Chongqing Municipal Health Commission (2020); Health Commission of Guangdong Province (2020); Health Commission of Zhejiang Province (2020); Health Commission of Hunan Province (2020).

| date | Beijing | Shanghai | Chongqing | Guangdong | Zhejiang | Hunan |
| --- | --- | --- | --- | --- | --- | --- |
| 2020.01.22 | 0 | 0 | 0 | 0 | 1 | 0 |
| 2020.01.23 | 0 | 0 | 0 | 2 | 1 | 0 |
| 2020.01.24 | 1 | 1 | 0 | 2 | 1 | 0 |
| 2020.01.25 | 2 | 1 | 0 | 2 | 1 | 0 |
| 2020.01.26 | 2 | 1 | 0 | 2 | 1 | 0 |
| 2020.01.27 | 2 | 3 | 0 | 4 | 3 | 0 |
| 2020.01.28 | 2 | 4 | 0 | 5 | 3 | 0 |
| 2020.01.29 | 4 | 5 | 0 | 6 | 4 | 0 |
| 2020.01.30 | 5 | 5 | 1 | 11 | 9 | 2 |
| 2020.01.31 | 5 | 9 | 1 | 12 | 15 | 3 |
| 2020.02.01 | 5 | 10 | 3 | 12 | 23 | 8 |
| 2020.02.02 | 9 | 10 | 7 | 14 | 36 | 16 |
| 2020.02.03 | 12 | 10 | 9 | 20 | 48 | 22 |
| 2020.02.04 | 24 | 15 | 14 | 32 | 63 | 35 |
| 2020.02.05 | 31 | 15 | 15 | 49 | 81 | 56 |
| 2020.02.06 | 33 | 25 | 24 | 52 | 98 | 91 |
| 2020.02.07 | 34 | 30 | 31 | 97 | 127 | 119 |
| 2020.02.08 | 37 | 41 | 39 | 125 | 173 | 159 |
| 2020.02.09 | 37 | 44 | 51 | 143 | 201 | 186 |
| 2020.02.10 | 48 | 48 | 66 | 181 | 250 | 213 |
| 2020.02.11 | 56 | 53 | 79 | 241 | 279 | 263 |
| 2020.02.12 | 68 | 57 | 102 | 284 | 327 | 312 |
| 2020.02.13 | 79 | 62 | 128 | 332 | 367 | 352 |
| 2020.02.14 | 97 | 90 | 152 | 386 | 409 | 389 |
| 2020.02.15 | 105 | 124 | 184 | 436 | 437 | 430 |
| 2020.02.16 | 114 | 140 | 207 | 473 | 470 | 467 |
| 2020.02.17 | 122 | 161 | 225 | 530 | 514 | 501 |
| 2020.02.18 | 145 | 177 | 254 | 571 | 544 | 542 |
| 2020.02.19 | 153 | 186 | 274 | 619 | 609 | 578 |
| 2020.02.20 | 169 | 199 | 299 | 664 | 647 | 638 |
| 2020.02.21 | 178 | 221 | 316 | 720 | 694 | 670 |

**Table 7:** Permanent population. The data for the six regions are available at China National Bureau of Statistics (2018).

| City/Province | Permanent population |
| --- | --- |
| Beijing | 21,540,000 |
| Shanghai | 24,240,000 |
| Guangdong | 113,460,000 |
| Chongqing | 31,020,000 |
| Hunan | 68,990,000 |
| Zhejiang | 57,370,000 |

## Appendix B: Formal Definition of the Markov Process

In this appendix, we present the transition rates of the system *ξ*(*t*) = *{S*(*t*), *E*_1_(*t*), *E*_2_(*t*), *E*_*q*_(*t*), *A*_1_(*t*), *A*_2_(*t*), *A*_*q*_(*t*), *IN*_1_(*t*), *IN*_2_(*t*), *IH*_*L*_(*t*), *IH*_*S*_(*t*), *R*_*A*_(*t*), *R*_*N*_ (*t*), *R*_*H*_ (*t*), *D*(*t*)}, which uniquely determines the continuous time Markov Process in our proposed model.

1. infection:
  - [*S, E*_1_, …, *D*] → [*S* − 1, *E*_1_ + 1, …, *D*] at rate 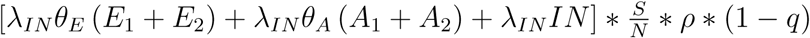.
  - [*S, E*_1_, *E*_2_…, *R, D*] → [*S* − 1, *E*_1_, *E*_2_ + 1…, *R, D*] at rate 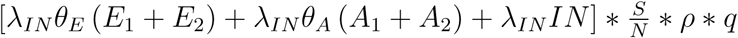.
  - [*S*, …, *A*_1_, …, *R, D*] → [*S* − 1, …, *A*_1_ + 1, …, *R, D*] at rate 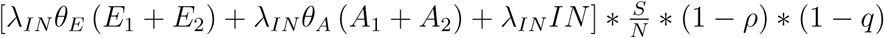
  - [*S*, …, *A*_2_, …, *R, D*] → [*S* − 1, …, *A*_2_ + 1, …, *R, D*] at rate 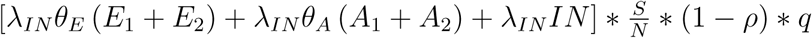
2. quarantine:
  - [*S, E*_1_, *E*_2_, *E*_*q*_…, *D*] → [*S, E*_1_, *E*_2_ *−* 1, *E*_*q*_ + 1…, *D*]at rate *E*_2_ *\* r*_*q*_.
  - [*S*, …, *A*_1_, *A*_2_, *A*_*q*_…, *D*] → [*S*, …, *A*_1_, *A*_2_ *−* 1, *A*_*q*_ + 1…, *D*]at rate *A*_2_ *\* r*_*q*_.
  - [*S*, …, *IN*_2_, …, *IH*_*L*_, …, *D*] → [*S*, …, *IN*_2_ *−* 1, …, *IH*_*L*_ + 1, …, *D*]at rate *IN*_2_*\*r*_*q*_ *\*p*_1_.
  - [*S*, …, *IN*_2_, …, *IH*_*S*_, …, *D*] → [*S*, …, *IN*_2_ *−* 1, …, *IH*_*S*_ + 1, …, *D*]at rate *IN*_2_ *\* r*_*q*_ *\** (1 *− p*_1_).
3. producing symptoms:
  - [*S, E*_1_, …, *IN*_1_, …, *D*] → [*S, E*_1_ *−* 1, …, *IN*_1_ + 1, …, *D*]at rate *E*_1_ *\* r*_*s*_.
  - [*S, E*_1_, *E*_2_…, *IN*_2_, …, *D*] → [*S, E*_1_, *E*_2_ *−* 1, …, *IN*_2_ + 1, …, *D*]at rate *E*_2_ *\* r*_*s*_.
  - [*S, E*_1_, *E*_2_, *E*_*q*_, …, *IH*_*L*_, …, *D*] → [*S, E*_1_, *E*_2_, *E*_*q*_ *−* 1, …, *IH*_*L*_ + 1, …, *D*]at rate *E*_*q*_ *\* r*_*s*_ *\* p*_1_.
  - [*S, E*_1_, *E*_2_, *E*_*q*_, …, *IH*_*S*_, …, *D*] → [*S, E*_1_, *E*_2_, *E*_*q*_ *−* 1, …, *IH*_*S*_ + 1, …, *D*]at rate *E*_*q*_ *\* r*_*s*_ *\** (1 *− p*_1_).
4. hospitalization:
  - [*S*, …, *IN*_1_, …, *IH*_*L*_, …, *D*] → [*S*, …, *IN*_1_ *−* 1, …, *IH*_*L*_ + 1, …, *D*] at rate *IN*_1_ *\*r*_*H*_ *\* q*_1_.
  - [*S*, …, *IN*_1_, …, *IH*_*S*_, …, *D*] → [*S*, …, *IN*_1_ *−* 1, …, *IH*_*S*_ + 1, …, *D*] at rate *IN*_1_ *\* r*_*H*_ *\** (1 *− q*_1_).
  - [*S*, …, *IN*_2_, …, *IH*_*L*_, …, *D*] → [*S*, …, *IN*_2_ *−* 1, …, *IH*_*L*_ + 1, …, *D*] at rate *IN*_2_ *\*r*_*H*_ *\* q*_1_.
  - [*S*, …, *IN*_2_, …, *IH*_*S*_, …, *D*] → [*S*, …, *IN*_2_ *−* 1, …, *IH*_*S*_ + 1, …, *D*] at rate *IN*_2_ *\* r*_*H*_ *\** (1 *− q*_1_).
5. symptom relief:
  - [*S*, …, *IH*_*L*_, *IH*_*S*_, …, *D*] → [*S*, …, *IH*_*L*_ + 1, *IH*_*S*_ *−* 1, …, *D*]at rate *IH*_*S*_ *\* r*_*b*_.
6. recovery:
  - [*S*, …, *A*_1_, …, *R*_*A*_, *R*_*N*_, *R*_*H*_, *D*] → [*S*, …, *A*_1_ *−* 1, …, *R*_*A*_ + 1, *R*_*N*_, *R*_*H*_, *D*] at rate *A*_1_ *\* γ*_*A*_.
  - [*S*, …, *A*_2_, …, *R*_*A*_, *R*_*N*_, *R*_*H*_, *D*] → [*S*, …, *A*_2_ *−* 1, …, *R*_*A*_ + 1, *R*_*N*_, *R*_*H*_, *D*] at rate *A*_2_ *\* γ*_*A*_.
  - [*S*, …, *IN*_1_, …, *R*_*A*_, *R*_*N*_, *R*_*H*_, *D*] → [*S*, …, *IN*_1_ *−* 1, …, *R*_*A*_, *R*_*N*_ + 1, *R*_*H*_, *D*] at rate *IN*_1_ *\* γ*_*I*_*N*.
  - [*S*, …, *IN*_2_, …, *R*_*A*_, *R*_*N*_, *R*_*H*_, *D*] → [*S*, …, *IN*_2_ *−* 1, …, *R*_*A*_, *R*_*N*_ + 1, *R*_*H*_, *D*] at rate *IN*_2_ *\* γ*_*I*_*N*.
  - [*S*, …, *IH*_*L*_, …, *R*_*A*_, *R*_*N*_, *R*_*H*_, *D*] → [*S*, …, *IH*_*L*_ *−* 1, …, *R*_*A*_, *R*_*N*_, *R*_*H*_ + 1, *D*] at rate *IH*_*L*_ *\* γ*_*I*_*H*.
7. death:
  - [*S*, …, *IN*_1_, …, *D*] → [*S*, …, *IN*_1_ *−* 1, ‥, *D* + 1]at rate *IN*_1_ *\* δ*_*IN*_.
  - [*S*, …, *IN*_2_, …, *D*] → [*S*, …, *IN*_2_ *−* 1, ‥, *D* + 1]at rate *IN*_2_ *\* δ*_*IN*_.
  - [*S*, …, *IH*_*S*_, …, *D*] → [*S*, …, *IH*_*S*_ *−* 1, ‥, *D* + 1]at rate *IH*_*S*_ *\* δ*_*IH*_.

